# Racial and Ethnic Disparities in Risk of Cardiovascular Disease in Women Treated for Breast Cancer

**DOI:** 10.64898/2026.04.23.26351612

**Authors:** Song Yao, Alexa Zimbalist, Haiyang Sheng, Peter Fiorica, Richard K. Cheng, Lucas Mendicino, Angela R. Omilian, Qianqian Zhu, Janise M. Roh, Cecile A. Laurent, Valerie S. Lee, Isaac J. Ergas, Carlos Iribarren, Jamal S. Rana, Mai Nguyen-Huynh, Eileen Rillamas-Sun, Dawn L. Hershman, Christine B. Ambrosone, Lawrence H. Kushi, Heather Greenlee, Marilyn L. Kwan

**Affiliations:** Department of Cancer Prevention and Control, Roswell Park Comprehensive Cancer Center, Buffalo, NY; Division of Research, Kaiser Permanente Northern California, Pleasanton, CA; University of Washington School of Medicine, Seattle, WA; Fred Hutchinson Cancer Center, Seattle, WA; Department of Biostatistics and Bioinformatics, Roswell Park Comprehensive Cancer Center,Buffalo, NY; Oakland Medical Center, Kaiser Permanente Northern California, Oakland, CA; Walnut Creek Medical Center, Kaiser Permanente Northern California, Walnut Creek, CA; Herbert Irving Comprehensive Cancer Center, Columbia University Irving Medical Center, New York, NY

**Keywords:** CVD, breast cancer, racial disparities, genetic ancestry

## Abstract

Racial and ethnic disparities exist in cardiovascular disease (CVD) burden in the general population; yet surprisingly few studies have examined such disparities in breast cancer patients, who are at higher risk due to cardiotoxic therapy. To investigate incidence of CVD and cardiometabolic risk factors across Asian, non-Hispanic Black (NHB), Hispanic, and non-Hispanic White (NHW) women with a history of breast cancer. In 4,071 women with breast cancer from a prospective cohort, the incidence of cardiometabolic risk factors and CVD occurring after breast cancer diagnosis were analyzed with self-identified race and ethnicity (SIRE) and global genetic ancestry. Racial and ethnic differences existed in the prevalence of cardiometabolic risk factors and CVD before breast cancer diagnosis, which continued to manifest in incident cases after cancer treatment. Asian, NHB, and Hispanic women were all at higher risk of diabetes than NHW women. Nonetheless, only NHB women had higher risk of CVD events, and Hispanic women were at lower risk. The apparent lower risk of CVD in Asian women largely disappeared after adjustment for covariates. Similar differences across SIRE groups were found in the cardiotoxic chemotherapy subgroup and the subgroup without chemotherapy, except for any CVD and VTE showing modifying effects of cardiotoxic chemotherapy. Analyses of genetic ancestry revealed similar results to SIRE. Our study reveals racial and ethnic disparities in cardiometabolic risk factors and CVD events before and after breast cancer diagnosis. Clinical and research attention is warranted to bridge the population-level gaps in CVD morbidity and mortality.

**Statement of Significance:** Our study provides strong evidence for racial and ethnic disparities in cardiovascular disease before and after breast cancer diagnosis. Clinical and research attention is warranted to bridge these population-level gaps.

## Introduction

Women with a history of breast cancer are at increased risk of cardiovascular disease (CVD) and cardiometabolic risk factors due to the cardiotoxic effects of breast cancer therapies (1-4). As a result, CVD has become the most common noncancer cause of death in breast cancer survivors in the US (5).

In the general population, the risk of CVD and cardiometabolic risk factors varies by self-identified race and ethnicity (SIRE) (6,7). Non-Hispanic Black (NHB) women have higher incidence and mortality of CVD events compared with non-Hispanic White (NHW) women (8), and a higher prevalence of cardiometabolic risk factors, particularly hypertension (9). Despite a higher prevalence of diabetes, obesity, and metabolic syndrome, Hispanic/Latina women have paradoxically lower rates of CVD mortality than NHW women (9). Moreover, although Asian women have the highest prevalence of type 2 diabetes (10), their rates of CVD is lower than NHW women (6).

To date, surprisingly few studies have investigated disparities in cardiovascular health in women after cancer treatment, with the limited literature suggesting more frequent cardiotoxicity from breast cancer therapy in NHB women than NHW women (11,12). One study also showed lower CVD mortality in Asian and Pacific Islander women with breast cancer than their NHW counterparts (13). These studies focused on only one racial and ethnic minority group compared with NHW women, and none included all major groups in the US in one integrative analysis. Further, few studies on racial and ethnic disparities of CVD have incorporated genetic ancestry data. While SIRE is a sociopolitical construct reflecting an individual’s lived experience, genetic ancestry is an alternative measure of human variations that reflects both biological and non-biological factors important to the causes of health disparities. In populations with admixed continental ancestries, including NHB and Hispanic groups in the US, the considerable variations in ancestral composition in an individual’s genetic background provide unique information relevant to health and disease status beyond what is captured by SIRE.

In the present analysis based on the Pathways Heart Study, we examined SIRE and genetic ancestry in relation to risk of cardiometabolic risk factors and CVD in a multi-ethnic cohort of breast cancer patients with long-time follow up.

## Methods

### Study population

The Pathways Study is a prospective cohort of women newly diagnosed with incident invasive breast cancer at Kaiser Permanente Northern California (KPNC) and enrolled between 2006 and 2013 with ongoing active follow up. The Pathways Heart Study was later established to investigate cardiometabolic risk factors and CVD events in women with and without a history of breast cancer. Details of the Pathways Study and the Pathways Heart Study have been published elsewhere (1,2,14,15). For this analysis, 4,071 breast cancer cases who were in both studies and had genotype data available were included. All patients have provided written informed consent, and this study was approved by Institutional Review Boards at all participating institutions.

### Data collection and measures

The main study endpoints were cardiometabolic risk factors (hypertension, diabetes, and dyslipidemia) and CVD events (arrhythmia, heart failure or cardiomyopathy, cardiac arrest, myocarditis or pericarditis, ischemic heart disease, transient ischemic attack (TIA), stroke, carotid disease, valvular disease, venous thromboembolism (VTE), and CVD-related death). The occurrence of cardiac arrest, myocarditis or pericarditis, TIA, and carotid disease was rare in this patient population and thus grouped for analysis. Arrhythmia, heart failure or cardiomyopathy, cardiac arrest, ischemic heart disease or stroke were combined into a composite event of serious CVD

Cardiometabolic risk factors were identified from the KPNC electronic health record (EHR) according to the Interaction Classification of Disease (ICD) diagnostic codes and Current Procedural Terminology (CPT) codes from primary care visits and hospitalization, in combination with abnormal laboratory results and use of indicative medications (2). CVD events were ascertained based on ICD codes and CPT codes from inpatient, ambulatory, and emergency department encounters, and/or hospital discharge records (1). For incident cardiometabolic risk factors identified by two or more diagnostic codes and/or medication dispensing record, the date associated with the first diagnosis code was considered the diagnosis date of the cardiometabolic risk factor. The time window used to identify incident cardiometabolic risk factors and CVD conditions was from the date of breast cancer diagnosis to December 31, 2021.

### Genetic ancestry estimation

Peripheral blood and saliva samples were collected at the time of baseline interview in the Pathways Study for genomic DNA. Genotyping was conducted using Illumina Multi-Ethnic Genotyping Array (16). Global genetic ancestries, including African, Amerindian, Asian, and European were estimated using unsupervised ADMIXTURE v1.3 (17), which measures the proportion of genetic components in each individual that are similar to reference populations (18,19).

### Statistical analysis

Descriptive statistics were used to summarize patient characteristics in the full cohort and by SIRE group - Asian, NHB, Hispanic, and NHW, which were self-identified at the time of baseline interview. Those self-identified as Native American, Alaska Native, other, or unknown race and ethnicity were not analyzed separately due to small numbers (total n=94). To examine whether racial and ethnic differences existed prior to breast cancer diagnosis, prevalent cardiometabolic risk factors and CVD were related to SIRE using logistic regression. Cumulative incidence curves for cardiometabolic risk factors and CVD events occurring after breast cancer diagnosis were plotted by SIRE, with p-values derived from log-rank test.

Fine and Gray hazards models were used to test the associations of SIRE with incident cardiometabolic risk factors and CVD events, with NHW as the referent group and all-cause death as competing risk (20). The time scale was defined as time since date of breast cancer diagnosis to first of incident event, health plan disenrollment, death, or last update of health status on December 31, 2021. Sub-distribution hazards ratio (sHR) and 95% confidence interval (CI) were derived with adjustment for age at diagnosis, body mass index (BMI), menopausal status, smoking status, primary care utilization, annual household income, educational attainment, cancer treatment (including cardiotoxic chemotherapy, radiation therapy, and endocrine therapy), any prevalent non-outcome cardiometabolic risk factors or CVD events. In addition, when modeling one specific event of interest, women with a history of that event were excluded. A similar approach was used to analyze genetic ancestry where the estimated proportions of African, Amerindian, and Asian ancestry were entered into the multivariable models, leaving the proportion of European ancestry out as the reference. To examine whether any racial and ethnic differences in disease incidence were modified by cardiotoxic chemotherapy, analyses were stratified by chemotherapy subgroup (no chemotherapy, non-cardiotoxic chemotherapy, and cardiotoxic chemotherapy), and an interaction term between SIRE and chemotherapy subgroup was added to the hazards models and tested using likelihood ratio statistics. All analyses were performed in R 4.2.0, with two-sided p≤0.05 considered statistically significant.

## Results

### Patient characteristics

Of the 4,071 breast cancer patients, 512 (12.6%) self-identified as Asian, 305 (7.5%) as NHB, 447 (11.0%) as Hispanic, 2,713 (66.6%) as NHW, and 94 (2.3%) as other or unknown race and ethnicity. There were notable differences across SIRE groups for many of the descriptive variables examined (**Table 1)**.

**Table 1.** Descriptive characteristics of patient population.

| Patient characteristics | Full cohort<br>(n=4,071)* | Asian (n=512) | Non-Hispanic<br>Black (n=305) | Hispanic (n=447) | Non-Hispanic<br>White (n=2,713) |
| --- | --- | --- | --- | --- | --- |
|  | mean ± SD | mean ± SD | mean ± SD | mean ± SD | mean ± SD |
| Length of KPNC membership,<br>months | 217 ± 65 | 213 ± 67 | 203 ± 75 | 206 ± 71 | 222 ± 62 |
| Age at baseline, years | 59.6 ± 12.1 | 53.2 ± 11.1 | 57.0 ± 11.4 | 55.1 ± 12.1 | 61.9 ± 11.6 |
| BMI at baseline, kg/m2 | 28.7 ± 6.7 | 25.8 ± 5.0 | 32.8 ± 8.0 | 29.7 ± 6.4 | 28.6 ± 6.7 |
| Primary care visit utilization | 3.1 ± 3.4 | 2.7 ± 2.6 | 3.5 ± 3.4 | 3.2 ± 3.4 | 3.1 ± 3.6 |
|  | No. (%) | No. (%) | No. (%) | No. (%) | No. (%) |
| Age at diagnosis, years |  |  |  |  |  |
| <40 | 181 (4.4) | 54 (11) | 17 (5.6) | 35 (7.8) | 71 (2.6) |
| 40-49 | 674 (17) | 141 (28) | 62 (20) | 128 (29) | 321 (12) |
| 50-59 | 1,167 (29) | 168 (33) | 99 (32) | 143 (32) | 734 (27) |
| 60-69 | 1,189 (29) | 108 (21) | 85 (28) | 78 (17) | 891 (33) |
| ≥70 | 860 (21) | 41 (8.0) | 42 (14) | 63 (14) | 696 (26) |
| Postmenopausal status | 2,901 (71) | 274 (54) | 195 (64) | 255 (57) | 2,113 (78) |
| BMI at baseline, kg/m2 |  |  |  |  |  |
| Normal or underweight | 1,339 (33) | 258 (51) | 45 (15) | 105 (23) | 910 (34) |
| Overweight | 1,292 (32) | 175 (34) | 78 (26) | 158 (35) | 848 (31) |
| Obese | 1,426 (35) | 76 (15) | 180 (59) | 184 (41) | 948 (35) |
| Smoking status |  |  |  |  |  |
| Never smoker | 2,294 (57) | 421 (82) | 154 (51) | 299 (67) | 1,367 (51) |
| Former smoker | 1,566 (39) | 76 (15) | 120 (40) | 134 (30) | 1,202 (44) |
| Current smoker | 197 (4.9) | 14 (2.7) | 29 (9.6) | 12 (2.7) | 135 (5.0) |
| Educational attainment |  |  |  |  |  |
| High school or less | 619 (15) | 55 (11) | 53 (17) | 150 (34) | 344 (13) |
| Some college | 1,415 (35) | 95 (19) | 134 (44) | 182 (41) | 957 (35) |
| College graduate | 1,134 (28) | 248 (49) | 72 (24) | 73 (16) | 723 (27) |
| Post-graduate | 895 (22) | 113 (22) | 44 (15) | 42 (9.4) | 684 (25) |
| Household income |  |  |  |  |  |
| <\$25K | 384 (9.4) | 48 (9.4) | 42 (14) | 53 (12) | 236 (8.7) |
| \$25K-49K | 752 (18) | 68 (13) | 66 (22) | 89 (20) | 506 (19) |
| \$50K-89K | 1,168 (29) | 129 (25) | 95 (31) | 145 (32) | 770 (28) |
| ≥\$90K | 1,304 (32) | 215 (42) | 57 (19) | 88 (20) | 914 (34) |
| Unknown | 463 (11) | 52 (10) | 45 (15) | 72 (16) | 287 (11) |
| AJCC stage |  |  |  |  |  |
| I | 2,219 (55) | 273 (53) | 143 (47) | 238 (53) | 1,525 (56) |
| II | 1,399 (34) | 191 (37) | 114 (37) | 154 (34) | 896 (33) |
| III | 395 (9.7) | 44 (8.6) | 40 (13) | 46 (10) | 256 (9.4) |
| IV | 58 (1.4) | 4 (0.8) | 8 (2.6) | 9 (2.0) | 36 (1.3) |
| Tumor ER negativity | 667 (16) | 85 (17) | 106 (35) | 83 (19) | 370 (14) |
| Tumor HER2 positivity | 522 (13) | 91 (18) | 46 (16) | 64 (15) | 307 (12) |
| Breast cancer treatment |  |  |  |  |  |
| Had mastectomy | 1,527 (38) | 250 (49) | 104 (34) | 183 (41) | 947 (35) |
| Had chemotherapy | 2,056 (51) | 294 (57) | 179 (59) | 251 (56) | 1,273 (47) |
| Had anti-HER2 therapy | 417 (10) | 69 (13) | 40 (13) | 52 (12) | 245 (9) |
| Had radiation therapy | 2,743 (67) | 303 (59) | 213 (70) | 308 (69) | 1,860 (69) |
| Had endocrine therapy | 3,079 (76) | 398 (78) | 182 (60) | 330 (74) | 2,110 (78) |

**Footnote:** \*94 patients of other or unknown self-reported race and ethnicity was included in all cases but not as a separate group.

### Prevalent cardiometabolic risk factors and CVD

At the time of breast cancer diagnosis, marked differences existed across SIRE groups in the prevalence of cardiometabolic risk factors, which was the highest in NHB women (**Supplementary Figure S1A**). Multivariable regression models further show that NHB, Asian, and Hispanic women were all at higher risk of prevalent diabetes than NHW women (**Supplementary Tables S1A**). Interestingly, for any cardiometabolic risk factor, hypertension, and dyslipidemia, lower risk was initially seen in Asian than NHW women in the unadjusted models, which became higher risk after adjustment for covariates. Similar results were seen for any cardiometabolic risk factor and dyslipidemia in Hispanic women.

Significant differences in the prevalence of CVD were also present at the time of breast cancer diagnosis, where NHB women had the highest rate of CVD events, and Asian and Hispanic women had the lowest rate (**Supplementary Figure S1B**). The higher prevalences of any CVD events, heart failure or cardiomyopathy, and stroke in Black women were confirmed in multivariable models (**Supplementary Table S1B**). Similar to the prevalent cardiometabolic risk factors, the lower risk of CVD initially seen in Asian and Hispanic women became weaker and non-significant after adjustment for covariates. In Asian women, an increased odds of heart failure or cardiomyopathy and ischemic heart disease was observed.

### Incident cardiometabolic risk factor events after breast cancer diagnosis

**Figure 1A** illustrates the cumulative incidence of cardiometabolic risk factor events by SIRE, with 2-year and 10-year cumulative incidence rates (CIRs) shown in **Supplementary Tables S2A and S3A**, respective. Of all SIRE groups, NHB women had the highest cumulative incidence of hypertension (10-year CIR: 37.2%) and diabetes (16.4%), and the incidence of diabetes was also higher in Asian (14.0%) and Hispanic (15.3%) women than in NHW women (7.7%). In multivariable models (**Table 2A**), all non-White groups had higher risk of diabetes than NHW women: Asian (sHR=2.99, 95% CI 2.20-4.05), NHB (sHR=1.78, 95% CI 1.28-2.48), and Hispanic (sHR=2.18, 95% CI 1.63-2.91) women. Moreover, NHB women were at higher risk of any cardiometabolic risk factor and hypertension; and Asian women at higher risk of any cardiometabolic risk factor, hypertension and dyslipidemia, which appeared only in the adjusted models, with the changes in risk estimates driven by younger age at diagnosis, lower BMI, and lower prevalence of smoking history in Asian women relative to NHW women.

**Table 2.** Associations of self-reported race and ethnicity with risk of incident cardiometabolic risk factor and cardiovascular disease events after breast cancer treatment.

| Incident event | Asian, self-identified |  | Non-Hispanic Black, self-identified |  | Hispanic, self-identified |  |
| --- | --- | --- | --- | --- | --- | --- |
|  | Unadjusted sHR (95% CI) | Adjusted sHR (95% CI) | Unadjusted sHR (95% CI) | Adjusted sHR (95% CI) | Unadjusted sHR (95% CI) | Adjusted sHR (95% CI) |
| **2A. Cardiometabolic risk factors (CMD)** |  |  |  |  |  |  |
| Any CMD risk factor | 0.98 (0.80, 1.20) | **1.53 (1.23, 1.91)** | 1.24 (0.94, 1.63) | **1.39 (1.04, 1.86)** | 0.89 (0.72, 1.09) | 0.95 (0.75, 1.20) |
| Hypertension | 0.86 (0.69, 1.09) | **1.30 (1.01, 1.67)** | **1.53 (1.15, 2.03)** | **1.72 (1.28, 2.32)** | 0.88 (0.70, 1.12) | 0.96 (0.75, 1.24) |
| Diabetes | 1.93 (1.48, 2.51) | **2.99 (2.20, 4.05)** | **2.35 (1.73, 3.18)** | **1.78 (1.28, 2.48)** | **2.08 (1.60, 2.72)** | **2.18 (1.63, 2.91)** |
| Dyslipidemia | 1.05 (0.86, 1.29) | **1.50 (1.20, 1.88)** | 0.97 (0.74, 1.27) | 1.03 (0.78, 1.37) | 0.99 (0.80, 1.24) | 1.12 (0.88, 1.41) |
| **2B. Cardiovascular diseases (CVD)** |  |  |  |  |  |  |
| Any CVD | **0.52 (0.40, 0.67)** | 0.81 (0.61, 1.07) | **1.29 (1.01, 1.64)** | **1.33 (1.03, 1.71)** | **0.51 (0.39, 0.68)** | **0.64 (0.48, 0.86)** |
| Serious CVD | **0.53 (0.39, 0.70)** | 0.85 (0.63, 1.16) | 1.22 (0.93, 1.60) | **1.35 (1.02, 1.79)** | **0.45 (0.32, 0.62)** | **0.61 (0.43, 0.85)** |
| Arrhythmia | **0.32 (0.21, 0.50)** | **0.59 (0.37, 0.92)** | 0.99 (0.71, 1.39) | 1.12 (0.79, 1.60) | **0.25 (0.15, 0.42)** | **0.33 (0.19, 0.56)** |
| Heart failure or cardiomyopathy | **0.47 (0.28, 0.77)** | 1.00 (0.59, 1.69) | **1.76 (1.23, 2.52)** | **1.60 (1.08, 2.36)** | **0.34 (0.19, 0.63)** | **0.45 (0.24, 0.83)** |
| Ischemic heart disease | 1.01 (0.65, 1.56) | 1.59 (0.98, 2.58) | 1.56 (0.98, 2.48) | **1.70 (1.04, 2.77)** | **0.38 (0.19, 0.78)** | **0.44 (0.21, 0.92)** |
| Stroke | 0.65 (0.35, 1.22) | 1.20 (0.61, 2.39) | 1.03 (0.53, 1.97) | 1.17 (0.59, 2.31) | 0.67 (0.35, 1.30) | 0.83 (0.42, 1.65) |
| Venous thromboembolic disease | **0.46 (0.26, 0.81)** | 0.80 (0.44, 1.46) | **1.84 (1.23, 2.74)** | **1.65 (1.07, 2.54)** | 0.87 (0.55, 1.37) | 0.94 (0.58, 1.54) |
| CVD-related death | **0.40 (0.22, 0.74)** | 0.87 (0.45, 1.66) | 0.68 (0.37, 1.26) | 0.75 (0.40, 1.41) | **0.54 (0.31, 0.96)** | 0.80 (0.45, 1.44) |

**Footnote:** Fine and Gray hazard models were used to test the associations of self-identified race and ethnicity (SIRE) (non-Hispanic White group as reference) with incident cardiometabolic risk factors and CVD events, with consideration of competing risk from all-cause death. The time scale was defined as time since date of breast cancer diagnosis to first of incident event, health plan disenrollment, death, or end of study. Sub-distribution hazards ratio (sHR) and 95% confidence interval (CI) were derived with adjustment for age at diagnosis, baseline BMI, menopausal status, smoking status, primary care utilization, household income, educational attainment, cancer treatment (including cardiotoxic chemotherapy, radiation therapy, and endocrine therapy), any prevalent non-outcome cardiometabolic risk factors or CVD events. In addition, when modeling one specific event of interest, women with a history of that event were excluded.

**Figure 1.**
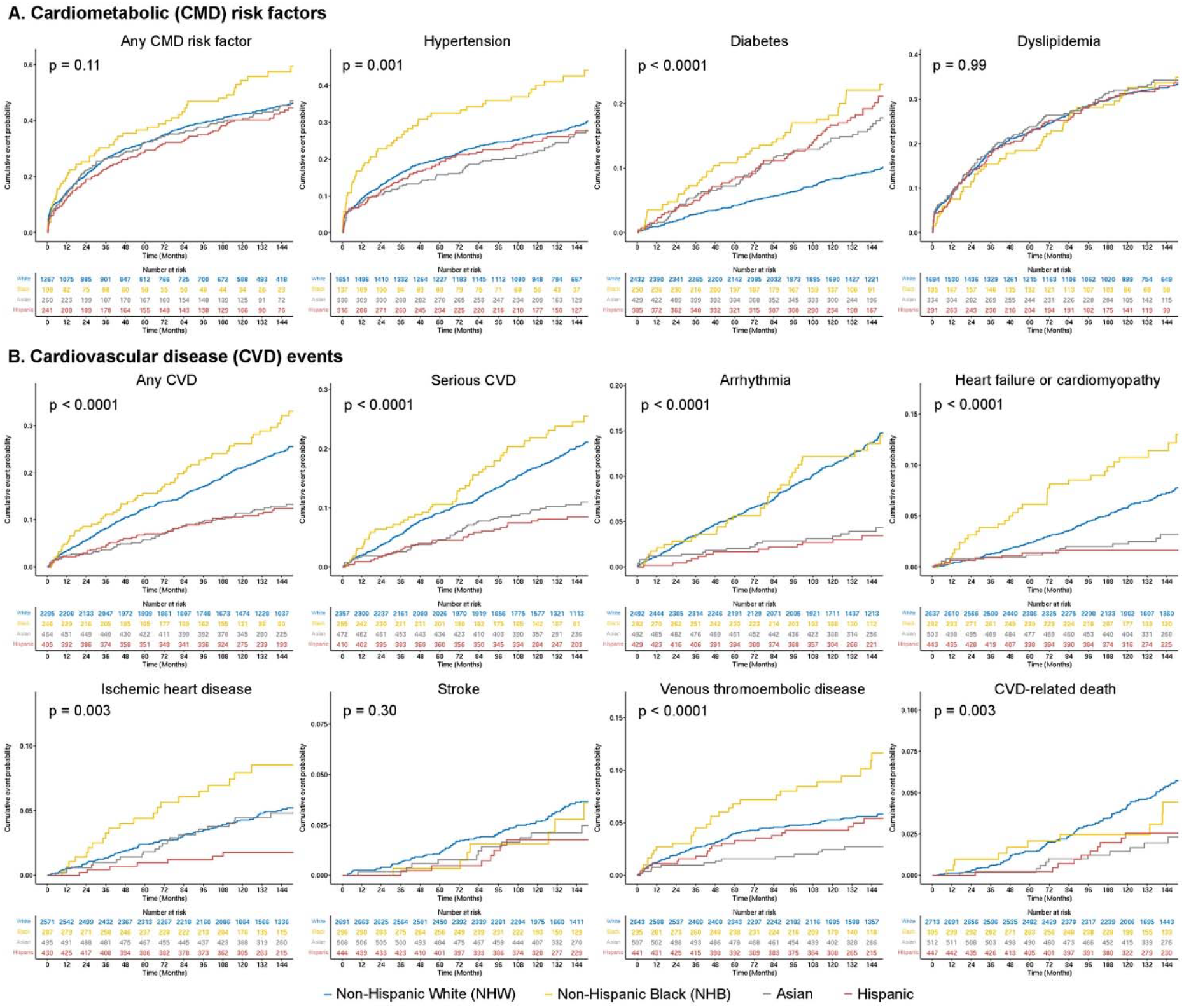
Cumulative incidence curves of cardiometabolic risk factors and cardiovascular disease by self-identified racial and ethnic groups Cumulative incidence on the Y-axis was plotted against time of follow up (months) on the x-axis by self-identified racial and ethnic group (SIRE) for each cardiometabolic risk factor **(A)** and cardiovascular disease event **(B)**. The time scale was defined as time since the date of breast cancer diagnosis to first of incident event, health plan disenrollment, death, or last update of health status on December 31, 2021. P-values were derived from log-rank test to compare the cumulative incidence across SIRE groups. Number of patients at risk at each year during follow up are shown at the bottom of the plots.

### Incident CVD events after breast cancer diagnosis

As shown in **Figure 1B** and **Supplementary Tables S2B and S3B**, NHB women had the highest cumulative incidence of any CVD (10-year CIR: 24.0%), followed by NHW women (20.2%), whereas Asian and Hispanic women had the lowest incidence (11.0% and 10.1%, respectively). Similar differences were noted for the incidence of serious CVD and heart failure or cardiomyopathy. The incidence of ischemic heart disease was the highest in NHB women and the lowest in Hispanic women, whereas NHW and Asian women had a similar incidence in between. The incidence of arrhythmia was similar between Asian and Hispanic women and lower than in NHB and NHW women. NHB women had the highest incidence of VTE and Asians had the lowest incidence, with NHW and Hispanic women sharing a similar incidence in between. Lastly, NHW women had the highest incidence of CVD-related death in all racial and ethnic groups.

In multivariable models (**Table 2B**), Asian women had lower risk of any CVD, serious CVD, arrhythmia, heart failure or cardiomyopathy, VTE, and CVD-related death; but all those associations, except for arrhythmia, became non-significant in the adjusted models. NHB women had higher risk of any CVD, serious CVD, heart failure or cardiomyopathy, ischemic heart disease, and VTE. There were no significant differences in CVD-related death between NHB and NHW women. In unadjusted models, Hispanic women had lower risk of any CVD, serious CVD, arrhythmia, heart failure or cardiomyopathy, ischemic heart disease, and CVD-related death than NHW women, all of which, except for CVD-related death, remained significant in the adjusted models.

### Impact of cardiotoxic chemotherapy

Of all the SIRE groups, NHB women had the highest rate (45.9%) of receiving cardiotoxic chemotherapy (anthracycline and/or trastuzumab) (**Supplementary Table S4**), followed by Hispanic (42.3%) and Asian (42.2%) women, whereas NHW women had the lowest rate (33.4%). **Supplementary Figures S2A** and **S2B** show the cumulative incidence of cardiometabolic risk factor and CVD events, respectively, across SIRE groups by the three chemotherapy subgroups (no chemotherapy, non-cardiotoxic chemotherapy, and cardiotoxic chemotherapy). Most of the racial and ethnic differences observed in the full cohort remained in similar patterns in the cardiotoxic chemotherapy subgroup, which included any cardiometabolic risk factor, hypertension, diabetes, any CVD, serious CVD, arrhythmia, heart failure or cardiomyopathy, ischemic heart disease, and VTE. The exception was CVD-related death, which showed no differences in this subgroup.

In the subgroup without chemotherapy, similar racial and ethnic differences to the full cohort were also found for diabetes, arrhythmia, heart failure or cardiomyopathy, and ischemic heart disease. However, the patterns appeared to differ between the no chemotherapy and the cardiotoxic chemotherapy subgroups for any cardiometabolic risk factor, any CVD, serious CVD, and VTE, all showing higher cumulative incidence in NHB women than NHW women in the cardiotoxic chemotherapy subgroup but not in the no chemotherapy subgroup. In addition, Hispanic women had lower incidence of VTE in the no chemotherapy subgroup, but no such difference was found in the cardiotoxic subgroup. Interaction testing and stratified analyses in multivariable models (**Table 3**) confirmed the modifying effects of chemotherapy subgroup on the associations of SIRE with risk of any CVD and VTE (P for interaction <0.001).

**Table 3.** Associations of self-identified race and ethnicity with incident any cardiovascular disease and venous thromboembolic event by chemotherapy subgroup.

| Chemotherapy subgroup | Asian, self-identified | Non-Hispanic Black, self-identified | Hispanic, self-identified | P for interaction |
| --- | --- | --- | --- | --- |
|  | sHR (95% CI) | sHR (95% CI) | sHR (95% CI) |  |
| Any cardiovascular disease (CVD) |  |  |  |  |
| No chemotherapy | **0.44 (0.29, 0.67)** | 1.06 (0.71, 1.57) | **0.46 (0.29, 0.71)** | <0.0001 |
| Non-cardiotoxic chemotherapy | 0.54 (0.27, 1.09) | 1.38 (0.63, 3.01) | 0.47 (0.20, 1.09) |  |
| Cardiotoxic chemotherapy | 0.60 (0.41, 0.88) | **1.49 (1.06, 2.11)** | **0.59 (0.40, 0.89)** |  |
| Venus thromboembolic event (VTE) |  |  |  |  |
| No chemotherapy | 0.49 (0.20, 1.21) | 1.73 (0.89, 3.36) | **0.33 (0.10, 1.05)** | 0.0002 |
| Non-cardiotoxic chemotherapy | 0.79 (0.23, 2.71) | 0.70 (0.09, 5.30) | 1.05 (0.31, 3.62) |  |
| Cardiotoxic chemotherapy | **0.33 (0.13, 0.81)** | **1.91 (1.13, 3.22)** | 1.13 (0.64, 1.98) |  |

**Footnote:** Fine and Gray hazard models were used to test the associations of self-identified race and ethnicity (SIRE) (non-Hispanic White group as reference) with incident any cardiovascular disease (CVD) and venous thromboembolic event (VTE), with consideration of competing risk from all-cause death. The analyses were stratified by chemotherapy subgroup (no chemotherapy, non-cardiotoxic chemotherapy, cardiotoxic chemotherapy). The time scale was defined as time since date of breast cancer diagnosis to first of incident event, health plan disenrollment, death, or end of study. Sub-distribution hazards ratio (sHR) and 95% confidence interval (CI) were derived with adjustment for age at diagnosis, baseline BMI, menopausal status, smoking status, primary care utilization, household income, educational attainment, cancer treatment (radiation therapy, and endocrine therapy), any prevalent non-outcome cardiometabolic risk factors or CVD events. In addition, when modeling one specific event of interest, women with a history of that event were excluded. P for interaction was derived from maximum likelihood test based on a base model without an interaction term between SIRE and chemotherapy subgroup and a full model with the interaction term.

### Genetic ancestry analysis

**Supplementary Table S5** summarizes the estimated global ancestry by SIRE group. There was little presence of non-European in NHW group and non-Asian ancestry in Asian group. In NHB and Hispanic groups, more than one major contributing ancestral components was identified. The median proportions of African and European ancestry in the NHB group were 0.82 (range: 0.30-1.00) and 0.17 (0-0.69), respectively. The median proportions of Amerindian, African, and European ancestry in the Hispanic group were 0.34 (0-0.68), 0.05 (0-0.13), and 0.58 (0.21-1.00), respectively.

In analysis with European ancestry as the reference, the associations of Asian ancestry with cardiometabolic risk factor (**Table 4A**) and CVD (**Table 4B**) events were consistent with those seen in self-reported Asian vs. NHW groups. Similarly, consistent results were found between African ancestry and NHB vs. NHW groups. For Amerindian ancestry, the associations were reminiscent of those seen with Hispanic vs. NHW groups, i.e., higher risk of diabetes and lower risk of any CVD, serious CVD, arrhythmia, heart failure or cardiomyopathy, and ischemic heart disease.

**Table 4.**
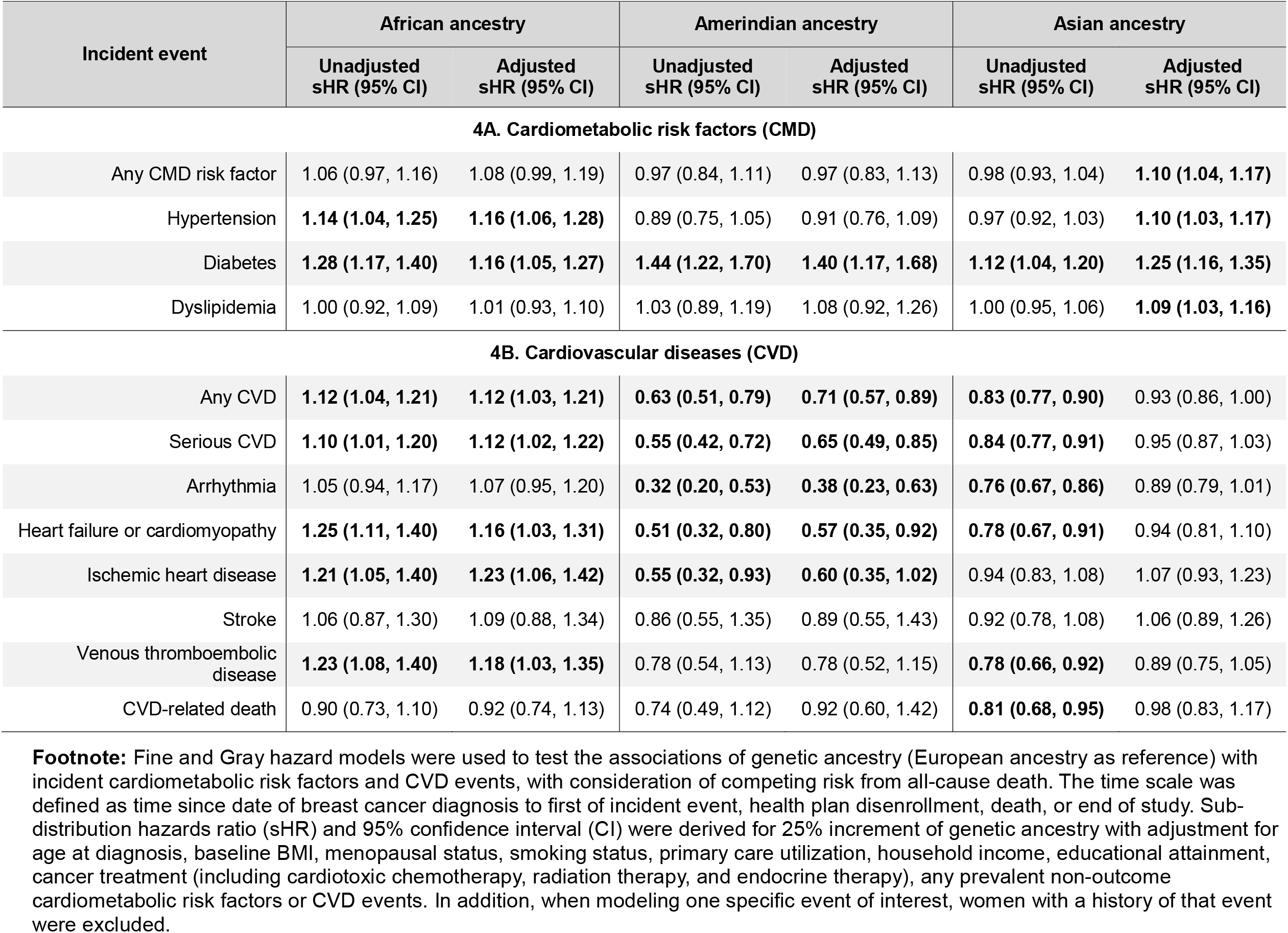
Associations of 25% increment of genetic ancestry with risk of incident cardiometabolic risk factor and cardiovascular disease events after breast cancer treatment.

| Incident event | African ancestry |  | Amerindian ancestry |  | Asian ancestry |  |
| --- | --- | --- | --- | --- | --- | --- |
|  | Unadjusted sHR (95% CI) | Adjusted sHR (95% CI) | Unadjusted sHR (95% CI) | Adjusted sHR (95% CI) | Unadjusted sHR (95% CI) | Adjusted sHR (95% CI) |
| **4A. Cardiometabolic risk factors (CMD)** |  |  |  |  |  |  |
| Any CMD risk factor | 1.06 (0.97, 1.16) | 1.08 (0.99, 1.19) | 0.97 (0.84, 1.11) | 0.97 (0.83, 1.13) | 0.98 (0.93, 1.04) | **1.10 (1.04, 1.17)** |
| Hypertension | **1.14 (1.04, 1.25)** | **1.16 (1.06, 1.28)** | 0.89 (0.75, 1.05) | 0.91 (0.76, 1.09) | 0.97 (0.92, 1.03) | **1.10 (1.03, 1.17)** |
| Diabetes | **1.28 (1.17, 1.40)** | **1.16 (1.05, 1.27)** | **1.44 (1.22, 1.70)** | **1.40 (1.17, 1.68)** | **1.12 (1.04, 1.20)** | **1.25 (1.16, 1.35)** |
| Dyslipidemia | 1.00 (0.92, 1.09) | 1.01 (0.93, 1.10) | 1.03 (0.89, 1.19) | 1.08 (0.92, 1.26) | 1.00 (0.95, 1.06) | **1.09 (1.03, 1.16)** |
| **4B. Cardiovascular diseases (CVD)** |  |  |  |  |  |  |
| Any CVD | **1.12 (1.04, 1.21)** | **1.12 (1.03, 1.21)** | **0.63 (0.51, 0.79)** | **0.71 (0.57, 0.89)** | **0.83 (0.77, 0.90)** | 0.93 (0.86, 1.00) |
| Serious CVD | **1.10 (1.01, 1.20)** | **1.12 (1.02, 1.22)** | **0.55 (0.42, 0.72)** | **0.65 (0.49, 0.85)** | **0.84 (0.77, 0.91)** | 0.95 (0.87, 1.03) |
| Arrhythmia | 1.05 (0.94, 1.17) | 1.07 (0.95, 1.20) | **0.32 (0.20, 0.53)** | **0.38 (0.23, 0.63)** | **0.76 (0.67, 0.86)** | 0.89 (0.79, 1.01) |
| Heart failure or cardiomyopathy | **1.25 (1.11, 1.40)** | **1.16 (1.03, 1.31)** | **0.51 (0.32, 0.80)** | **0.57 (0.35, 0.92)** | **0.78 (0.67, 0.91)** | 0.94 (0.81, 1.10) |
| Ischemic heart disease | **1.21 (1.05, 1.40)** | **1.23 (1.06, 1.42)** | **0.55 (0.32, 0.93)** | **0.60 (0.35, 1.02)** | 0.94 (0.83, 1.08) | 1.07 (0.93, 1.23) |
| Stroke | 1.06 (0.87, 1.30) | 1.09 (0.88, 1.34) | 0.86 (0.55, 1.35) | 0.89 (0.55, 1.43) | 0.92 (0.78, 1.08) | 1.06 (0.89, 1.26) |
| Venous thromboembolic disease | **1.23 (1.08, 1.40)** | **1.18 (1.03, 1.35)** | 0.78 (0.54, 1.13) | 0.78 (0.52, 1.15) | **0.78 (0.66, 0.92)** | 0.89 (0.75, 1.05) |
| CVD-related death | 0.90 (0.73, 1.10) | 0.92 (0.74, 1.13) | 0.74 (0.49, 1.12) | 0.92 (0.60, 1.42) | **0.81 (0.68, 0.95)** | 0.98 (0.83, 1.17) |

**Footnote:** Fine and Gray hazard models were used to test the associations of genetic ancestry (European ancestry as reference) with incident cardiometabolic risk factors and CVD events, with consideration of competing risk from all-cause death. The time scale was defined as time since date of breast cancer diagnosis to first of incident event, health plan disenrollment, death, or end of study. Sub-distribution hazards ratio (sHR) and 95% confidence interval (CI) were derived for 25% increment of genetic ancestry with adjustment for age at diagnosis, baseline BMI, menopausal status, smoking status, primary care utilization, household income, educational attainment, cancer treatment (including cardiotoxic chemotherapy, radiation therapy, and endocrine therapy), any prevalent non-outcome cardiometabolic risk factors or CVD events. In addition, when modeling one specific event of interest, women with a history of that event were excluded.

## Discussion

In a large contemporary cohort of women with a history of breast cancer, we found pronounced racial and ethnic differences in cardiometabolic risk factor and CVD events. Such disparities already existed at the time of breast cancer diagnosis and persisted after cancer treatment in incident cases, particularly in those receiving cardiotoxic chemotherapy.

Although the sample size of NHB patients in our study was relatively small, the findings of higher morbidity burden of CVD and cardiometabolic risk factors than NHW patients were clear and consistent with the prior literature (8,9,11,12). However, we did not observe higher CVD-related mortality in Black women, which conflicts with prior studies. Our limited sample size and number of CVD-related deaths might contribute to this unexpected finding. This could be due to survivor bias as NHB women might be more likely to die from breast cancer earlier during follow up. Although accounting for non-CVD-related death did not change the results, we cannot completely refute residual bias. Moreover, all patients in our study had the same insurance coverage through KPNC, and inequities in insurance might be one of the contributing factors to higher CVD mortality in Black women.

Despite having the highest prevalence of diabetes, self-identified Asian individuals tend to have lower burden of CVD morbidity and mortality than NHW individuals, including those with a history of breast cancer (13). Our data show higher morbidity of diabetes in Asian breast cancer patients. Intriguingly, the direction of the associations of Asian compared with NHW women with CVD and cardiometabolic risk factors flipped between the unadjusted and adjusted models, which was driven by younger age, lower BMI, and less smoking history in Asian than NHW women. Pending validation in future studies, our findings suggest that at similar age and BMI, Asian breast cancer patients might be at higher risk of multiple cardiometabolic conditions than NHW women, and their risk of CVD conditions might not be as low as previously believed.

Our data also demonstrate the phenomenon of the “Hispanic paradox” in CVD risk in breast cancer patients (9). Although Hispanic women had higher BMI, lower educational attainment, lower household income, and higher rates of diabetes than NHW women, they had significantly lower CVD risk, which remained after adjusting for covariates. The associations of Amerindian ancestry with diabetes and incident CVD were similar to the associations seen in Hispanic women. These results suggest that the lower risk of incident CVD events in Hispanic women might be driven, at least in part, by Amerindian ancestry.

We are unaware of prior reports of an association of Amerindian ancestry with lower CVD risk. On the contrary, self-identified American Indians in the Strong Heart Study, which included both men and women, had higher rates of coronary heart disease than any other racial or ethnic group, which were also more often fatal (21,22). In another study in Mexicans, Amerindian ancestry was associated with low HDL-cholesterol (23). The inconsistency might be due to breast cancer treatment, since the lower CVD risk in Hispanic women was more apparent for incident cases than for prevalent cases. If our findings are further validated in future studies, they will join the body of literature showing the contribution of Amerindian ancestry and/or its sociocultural influences to distinct risk profiles in Hispanic individuals, as previously demonstrated in the case of HER2-positive breast cancer and childhood acute lymphoblastic leukemia (24,25).

In the last two decades, the analysis of genetic ancestry has been commonly adopted in population genetics and health disparity research. Our study herein demonstrates that racial and ethnic differences exist in the morbidity of CVD and cardiometabolic risk factors in women after breast cancer treatment, whereby the results based on genetic ancestry were similar to those based on SIRE. Moreover, the findings of Amerindian ancestry being associated with lower risk of CVD morbidity that mirrored the association of Hispanic ethnicity suggest that genetic variants and/or social determinants more commonly found in Native American populations may be protective against the development of CVD, providing clues for future research. Such findings would have been overlooked without the use of genetic ancestry estimates.

One limitation of our study is the relatively limited sample size of non-White populations, particularly the NHB group, which could have led to suboptimal statistical power in some of the analyses. For those self-identified as Native American and Alaska Native, the sample sizes were even smaller that we could not reliably analyze as separate groups. More efforts are warranted to recruit the understudied populations in future prospective cohort studies like ours. Another limitation of our study comes from the challenge to distinguish between prevalent and incident diagnoses of CVD and cardiometabolic risk factors, as underdiagnosis of certain cardiometabolic or cardiovascular conditions was possible prior to breast cancer diagnosis, which then manifested as incident cases after breast cancer. We attempted to address this by excluding patients with less than 12 months of KP membership before breast cancer diagnosis and also excluded known prevalent cases in the analyses, but we could not completely be certain all prevalent diagnosis were accounted for.

In conclusion, our study provides strong evidence for racial and ethnic disparities in cardiometabolic risk factors and CVD events before and after breast cancer diagnosis. Clinical and research attention is warranted to bridge these population-level gaps in CVD risk.

## Data Availability

All data produced in the present study are available upon reasonable request to the authors.

## Abbreviations

(in alphabetical order)

BMI: body mass index
CI: confidence interval
CVD: cardiovascular disease
NHB: non-Hispanic Black
NHW: non-Hispanic White
sHR: subdistribution hazards ratio
SIRE: self-identified race and ethnicity
TIA: transient ischemic attack
VTE: venous thromboembolic event

## Acknowledgements

We thank the KPNC patients who provided the data for this study.

## Author Contributions

**Conception and design:** Song Yao, Heather Greenlee, Marilyn L. Kwan

**Financial support:** Song Yao, Marilyn L. Kwan, Heather Greenlee

**Administrative support:** Song Yao, Marilyn L. Kwan, Heather Greenlee, Lawrence H. Kushi, Christine B. Ambrosone

**Provision of study materials or patients:** Marilyn L. Kwan, Lawrence H. Kushi

**Collection and assembly of data:** Song Yao, Marilyn L. Kwan, Alexa Zimbalist, Cecile A. Laurent, Valerie S. Lee, Janise M. Roh, Heather Greenlee

**Data analysis and interpretation:** Song Yao, Marilyn L. Kwan, Alexa Zimbalist, Haiyang Sheng, Cecile A. Laurent, Janise M. Roh, Richard K. Cheng, Eileen Rillamas-Sun, Heather Greenlee

**Manuscript writing:** All authors

**Final approval of manuscript:** All authors

**Accountable for all aspects of the work:** All authors

