## Supplemental Tables 1-4 and Supplemental Figures 1-2 for "Racial and Ethnic Disparities in Risk of Cardiovascular Disease in Women Treated for Breast Cancer"

### Supplementary Materials

**Supplementary Table S1.** Association of self-reported race and ethnicity with risk of prevalent cardiometabolic risk factors and cardiovascular diseases at the time of breast cancer treatment

**Supplementary Table S2.** Estimated 2-year cumulative incidence rate and 95% confidence interval of cardiometabolic risk factor and cardiovascular disease events after breast cancer treatment

**Supplementary Table S3.** Estimated 10-year cumulative incidence rate and 95% confidence interval of cardiometabolic risk factor and cardiovascular disease events after breast cancer treatment

**Supplementary Table S4.** The rate of cardiotoxic chemotherapy treatment overall and by self-identified race and ethnicity

**Supplementary Table S5.** The proportion of estimated genetic ancestry by self-identified race and ethnicity

**Supplementary Figure S1.** Prevalence of cardiometabolic risk factors **(A)** and cardiovascular disease (CVD) **(B)** prior to breast cancer diagnosis by self-identified racial and ethnic groups

**Supplementary Figure S2.** Cumulative incidence of cardiometabolic risk factors **(A)** and cardiovascular disease (CVD) **(B)** by self-identified racial and ethnic groups in patients receiving no chemotherapy, non-cardiotoxic chemotherapy, and cardiotoxic chemotherapy

**Supplementary Table S1.** Association of self-reported race and ethnicity with risk of prevalent cardiometabolic risk factors and cardiovascular diseases at the time of breast cancer treatment

| Incident event | Asian, self-identified | | Non-Hispanic Black, self-identified | | Hispanic, self-identified | |
|---|---|---|---|---|---|---|
| | Unadjusted OR (95% CI) | Adjusted OR (95% CI) | Unadjusted OR (95% CI) | Adjusted OR (95% CI) | Unadjusted OR (95% CI) | Adjusted OR (95% CI) |
| **S1A. Cardiometabolic risk factors (CMD)** | | | | | | |
| Any CMD risk factor | 0.85 (0.70, 1.03) | **2.75 (2.15, 3.52)** | **1.60 (1.25, 2.05)** | **2.08 (1.54, 2.82)** | **0.75 (0.61, 0.92)** | 1.28 (0.99, 1.65) |
| Hypertension | **0.80 (0.66, 0.97)** | **2.55 (1.98, 3.29)** | **1.91 (1.50, 2.42)** | **2.66 (1.99, 3.58)** | **0.64 (0.52, 0.80)** | 0.97 (0.74, 1.27) |
| Diabetes | **1.67 (1.28, 2.17)** | **4.88 (3.49, 6.84)** | **1.90 (1.38, 2.60)** | **1.64 (1.14, 2.34)** | **1.39 (1.03, 1.86)** | **1.86 (1.31, 2.61)** |
| Dyslipidemia | 0.89 (0.73, 1.08) | **2.16 (1.70, 2.76)** | 1.08 (0.84, 1.37) | 1.25 (0.94, 1.65) | 0.89 (0.72, 1.10) | **1.45 (1.13, 1.86)** |
| **S1B. Cardiovascular diseases (CVD)** | | | | | | |
| Any CVD | **0.57 (0.41, 0.77)** | 1.22 (0.85, 1.73) | 1.32 (0.97, 1.77) | **1.67 (1.18, 2.33)** | **0.57 (0.40, 0.79)** | 0.80 (0.55, 1.14) |
| Arrhythmia | **0.46 (0.28, 0.71)** | 0.85 (0.52, 1.41) | 0.92 (0.57, 1.41) | 1.15 (0.70, 1.82) | **0.47 (0.28, 0.75)** | 0.62 (0.37, 1.05) |
| Heart failure or cardiomyopathy | 0.62 (0.29, 1.18) | **2.40 (1.09, 5.32)** | 1.54 (0.81, 2.72) | **1.99 (1.02, 3.88)** | **0.31 (0.10, 0.76)** | 0.49 (0.17, 1.43) |
| Ischemic heart disease | 0.62 (0.36, 1.01) | 1.75 (0.98, 3.11) | 1.14 (0.66, 1.83) | 1.38 (0.80, 2.39) | 0.72 (0.41, 1.16) | 1.10 (0.63, 1.92) |
| Stroke | 0.96 (0.28, 2.53) | 2.10 (0.56, 6.43) | **3.72 (1.61, 7.90)** | **4.77 (1.92, 11.10)** | 0.83 (0.20, 2.40) | 1.06 (0.24, 3.38) |
| Venous thromboembolic disease | 0.51 (0.18, 1.18) | 0.89 (0.33, 2.38) | 0.69 (0.21, 1.71) | 1.06 (0.37, 3.08) | 0.47 (0.14, 1.16) | 0.78 (0.27, 2.27) |

Footnote: Logistic regression models were used to relate prevalent cardiometabolic risk factors to self-reported race and ethnicity (SIRE), with non-Hispanic White group as reference. Odds ratio (OR) and 95% confidence interval (CI) were derived either with or without adjustment for age at diagnosis, baseline body mass index, menopausal status, smoking status, household income, and education levels.

**Supplementary Table S2.** Estimated 2-year cumulative incidence rate and 95% confidence interval of cardiometabolic risk factor and cardiovascular disease events after breast cancer treatment

| Incident event | Asian, self-identified | Non-Hispanic Black, self-identified | Hispanic, self-identified | Non-Hispanic White, self-identified |
|---|---|---|---|---|
| **S2A. Cardiometabolic risk factors** | | | | |
| Any CMD risk factor | 22.3 (17.7, 27.8) | 25.0 (17.8, 33.9) | 19.1 (14.6, 24.5) | 20.9 (18.8, 23.2) |
| Hypertension | 10.4 (7.5, 14.1) | 22.6 (16.4, 30.3) | 11.7 (8.6, 15.7) | 13.1 (11.5, 14.8) |
| Diabetes | 4.0 (2.5, 6.3) | 4.8 (2.8, 8.2) | 3.6 (2.2, 6.0) | 1.8 (1.4, 2.4) |
| Dyslipidemia | 14.7 (11.3, 18.9) | 11.4 (7.5, 16.7) | 14.1 (10.6, 18.6) | 13.5 (12.0, 15.2) |
| **S2B. Cardiovascular diseases (CVD)** | | | | |
| Any CVD | 2.8 (1.6, 4.7) | 8.5 (5.7, 12.7) | 3.0 (1.7, 5.1) | 5.7 (4.8, 6.7) |
| Serious CVD | 1.9 (1.0, 3.6) | 6.3 (3.9, 9.9) | 1.7 (0.8, 3.5) | 3.7 (3.0, 4.6) |
| Arrhythmia | 1.2 (0.6, 2.6) | 2.8 (1.4, 5.5) | 0.5 (0.1, 1.7) | 2.5 (1.9, 3.2) |
| Heart failure or cardiomyopathy | 0.8 (0.3, 2) | 3.1 (1.6, 5.8) | 0.7 (0.2, 2.0) | 0.8 (0.5, 1.2) |
| Ischemic heart disease | 0.6 (0.2, 1.8) | 1.4 (0.5, 3.5) | 0.2 (0, 1.3) | 0.9 (0.6, 1.3) |
| Stroke | 0.2 (0, 1.1) | 0 (0, 1.3) | 0 (0, 0.9) | 0.4 (0.2, 0.7) |
| Venous thromboembolic disease | 1 (0.4, 2.3) | 3.1 (1.6, 5.7) | 1.1 (0.5, 2.6) | 1.9 (1.5, 2.5) |
| CVD-related death | 0.2 (0, 1.1) | 1.0 (0.3, 2.9) | 0 (0, 0.9) | 0.3 (0.1, 0.6) |

**Supplementary Table S3.** Estimated 10-year cumulative incidence rate and 95% confidence interval of cardiometabolic risk factor and cardiovascular disease events after breast cancer treatment

| Incident event | Asian, self-identified | Non-Hispanic Black, self-identified | Hispanic, self-identified | Non-Hispanic White, self-identified |
|---|---|---|---|---|
| **S2A. Cardiometabolic risk factors** | | | | |
| Any CMD risk factor | 39.2 (33.5, 45.3) | 49.1 (39.8, 58.4) | 38.2 (32.3, 44.4) | 41.0 (38.4, 43.8) |
| Hypertension | 21.3 (17.3, 26.0) | 37.2 (29.6, 45.6) | 23.7 (19.4, 28.7) | 25.7 (23.6, 27.8) |
| Diabetes | 14.0 (11.0, 17.6) | 16.4 (12.3, 21.5) | 15.3 (12.1, 19.3) | 7.7 (6.7, 8.8) |
| Dyslipidemia | 31.1 (26.4, 36.3) | 29.2 (23.1, 36.1) | 29.6 (24.6, 35.0) | 29.9 (27.7, 32.1) |
| **S2B. Cardiovascular diseases (CVD)** | | | | |
| Any CVD | 11.0 (8.5, 14.2) | 24.0 (19.1, 29.7) | 10.1 (7.6, 13.4) | 20.2 (18.6, 21.9) |
| Serious CVD | 9.3 (7.0, 12.3) | 19.6 (15.2, 24.9) | 7.3 (5.2, 10.3) | 16.0 (14.6, 17.5) |
| Arrhythmia | 3.0 (1.9, 5.0) | 10.6 (7.6, 14.8) | 2.6 (1.4, 4.5) | 10.4 (9.3, 11.7) |
| Heart failure or cardiomyopathy | 2.2 (1.2, 3.9) | 9.6 (6.7, 13.5) | 1.6 (0.8, 3.2) | 5.3 (4.6, 6.3) |
| Ischemic heart disease | 4.2 (2.8, 6.4) | 7.0 (4.6, 10.5) | 1.6 (0.8, 3.3) | 4 (3.3, 4.8) |
| Stroke | 2.0 (1.1, 3.6) | 1.4 (0.5, 3.4) | 1.6 (0.8, 3.2) | 2.5 (1.9, 3.1) |
| Venous thromboembolic disease | 2.4 (1.4, 4.1) | 8.1 (5.5, 11.8) | 4.1 (2.6, 6.4) | 5.0 (4.2, 5.9) |
| CVD-related death | 1.4 (0.7, 2.8) | 2.3 (1.1, 4.7) | 2.2 (1.2, 4.1) | 3.8 (3.1, 4.6) |

**Supplementary Table S4.** The rate of cardiotoxic chemotherapy treatment overall and by self-identified race and ethnicity

| Self-identified race and ethnicity | No chemotherapy | Non-Cardiotoxic Chemotherapy | Cardiotoxic Chemotherapy |
|---|---|---|---|
| Full cohort (n=3,977) | 2051 (51.6%) | 474 (11.9%) | 1452 (36.5%) |
| Asian (n=512) | 224 (43.8%) | 72 (14.1%) | 216 (42.2%) |
| Non-Hispanic Black (n=305) | 132 (43.3%) | 33 (10.8%) | 140 (45.9%) |
| Hispanic (n=447) | 201 (45.0%) | 57 (12.8%) | 189 (42.3%) |
| Non-Hispanic White (n=2,713) | 1494 (55.1%) | 312 (11.5%) | 907 (33.4%) |

**Footnote:** Cardiotoxic chemotherapy includes anthracycline without trastuzumab, anthracycline with trastuzumab, and trastuzumab without anthracycline; non-cardiotoxic chemotherapy includes cyclophosphamide, fluoropyrimidine, and/or taxanes.

**Supplementary Table S5.** The proportion of estimated genetic ancestry by self-identified race and ethnicity

| Self-identified race and ethnicity | African ancestry | Amerindian ancestry | Asian ancestry | European ancestry |
|---|---|---|---|---|
| Asian | 0 (0-0.020) | 0 (0-0.046) | 0.981 (0.809-1.00) | 0 (0-0.182) |
| Non-Hispanic Black | 0.817 (0.297-1.00) | 0.007 (0-0.095) | 0.007 (0-0.027) | 0.173 (0-0.694) |
| Hispanic | 0.046 (0-0.130) | 0.338 (0-0.679) | 0.021 (0-0.513 | 0.577 (0.207-0.999) |
| Non-Hispanic White | 0.001 (0-0.100) | 0.002 (0-0.107) | 0.002 (0-0.115) | 0.991 (0.873-1.000) |

**Supplementary Figure S1.** Prevalence of cardiometabolic risk factors **(A)** and cardiovascular disease (CVD) **(B)** prior to breast cancer diagnosis by self-identified racial and ethnic groups

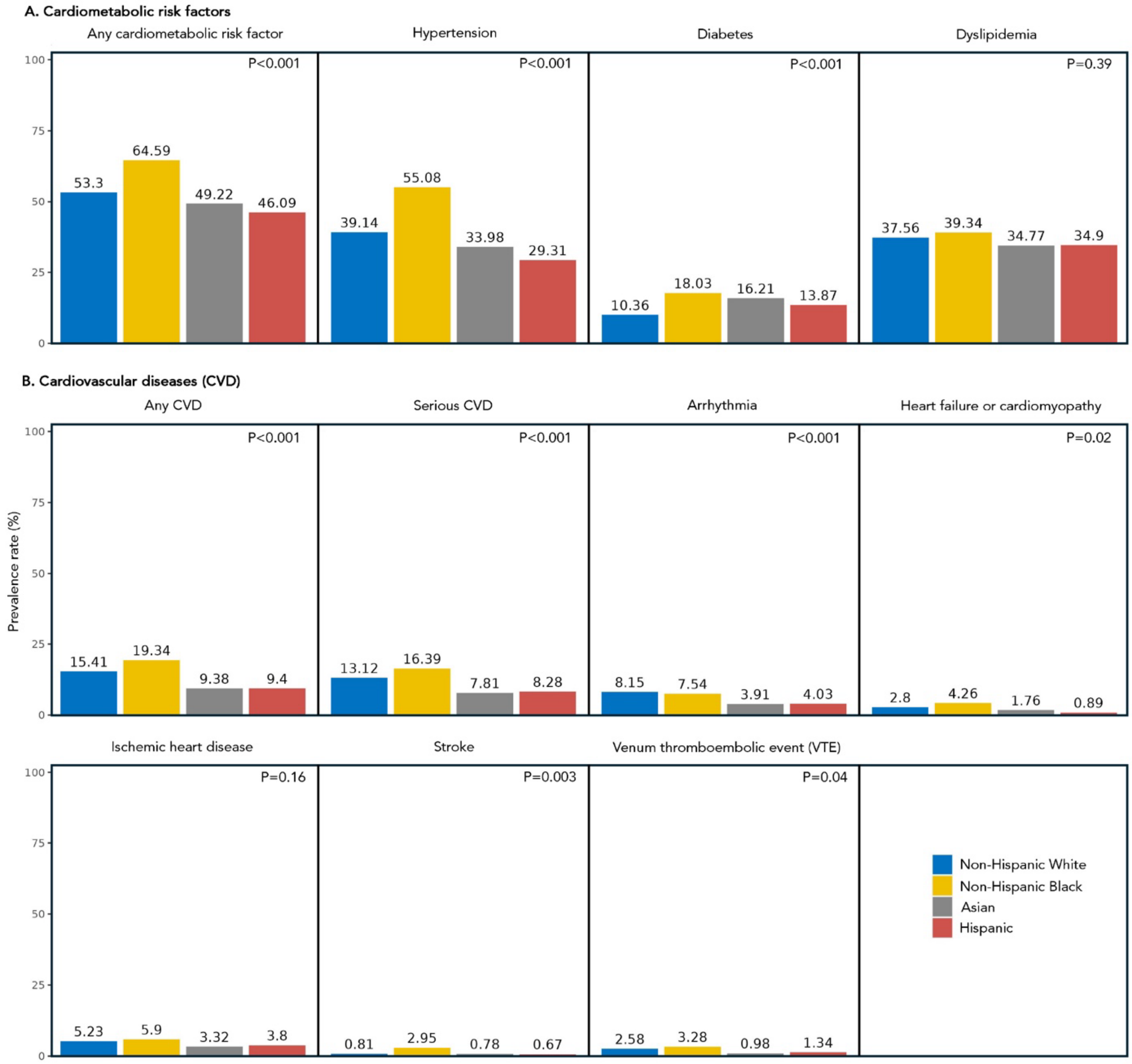

**Supplementary Figure S2.** Cumulative incidence of cardiometabolic risk factors **(A)** and cardiovascular disease (CVD) **(B)** by self-identified racial and ethnic groups in patients receiving no chemotherapy, non-cardiotoxic chemotherapy, and cardiotoxic chemotherapy

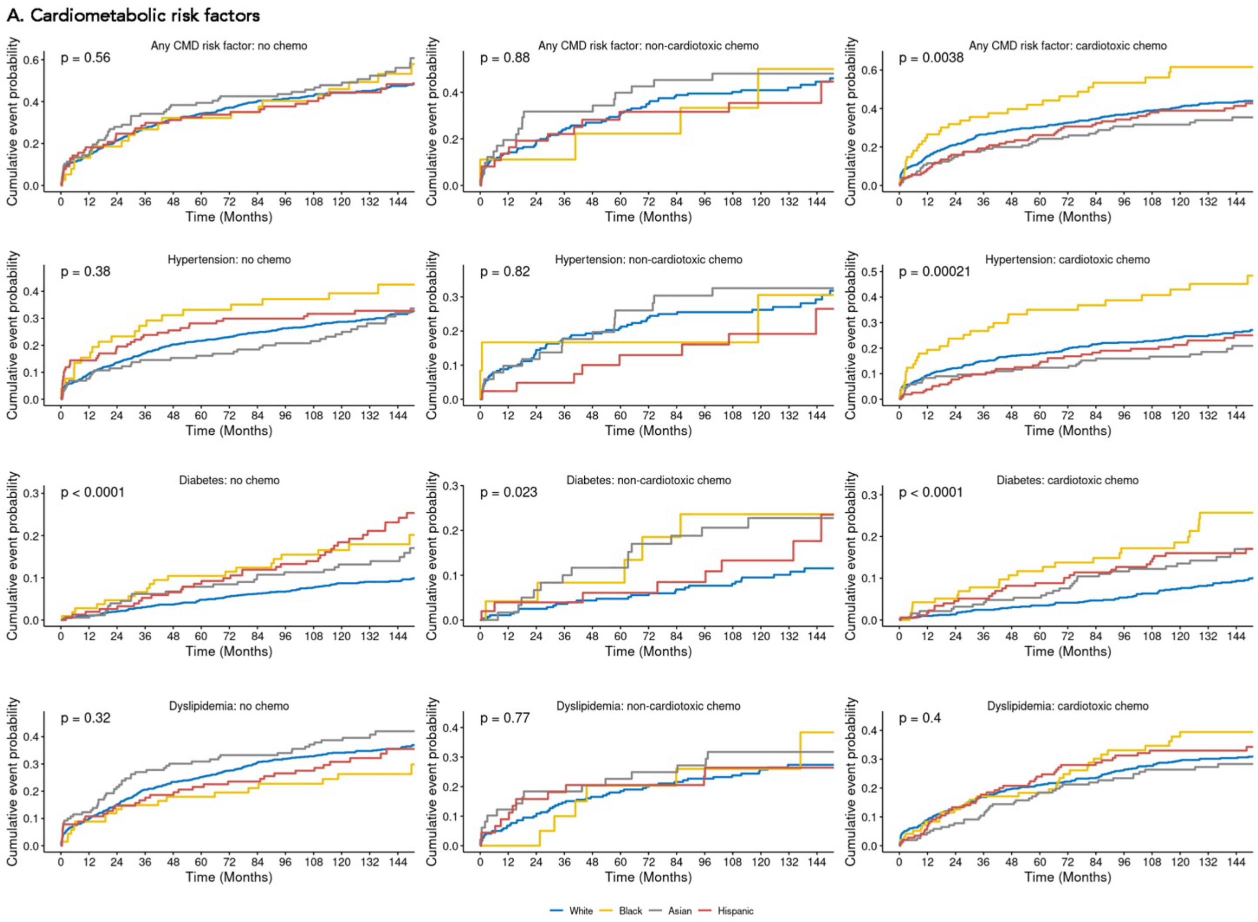

### B. Cardiovascular diseases (CVD)

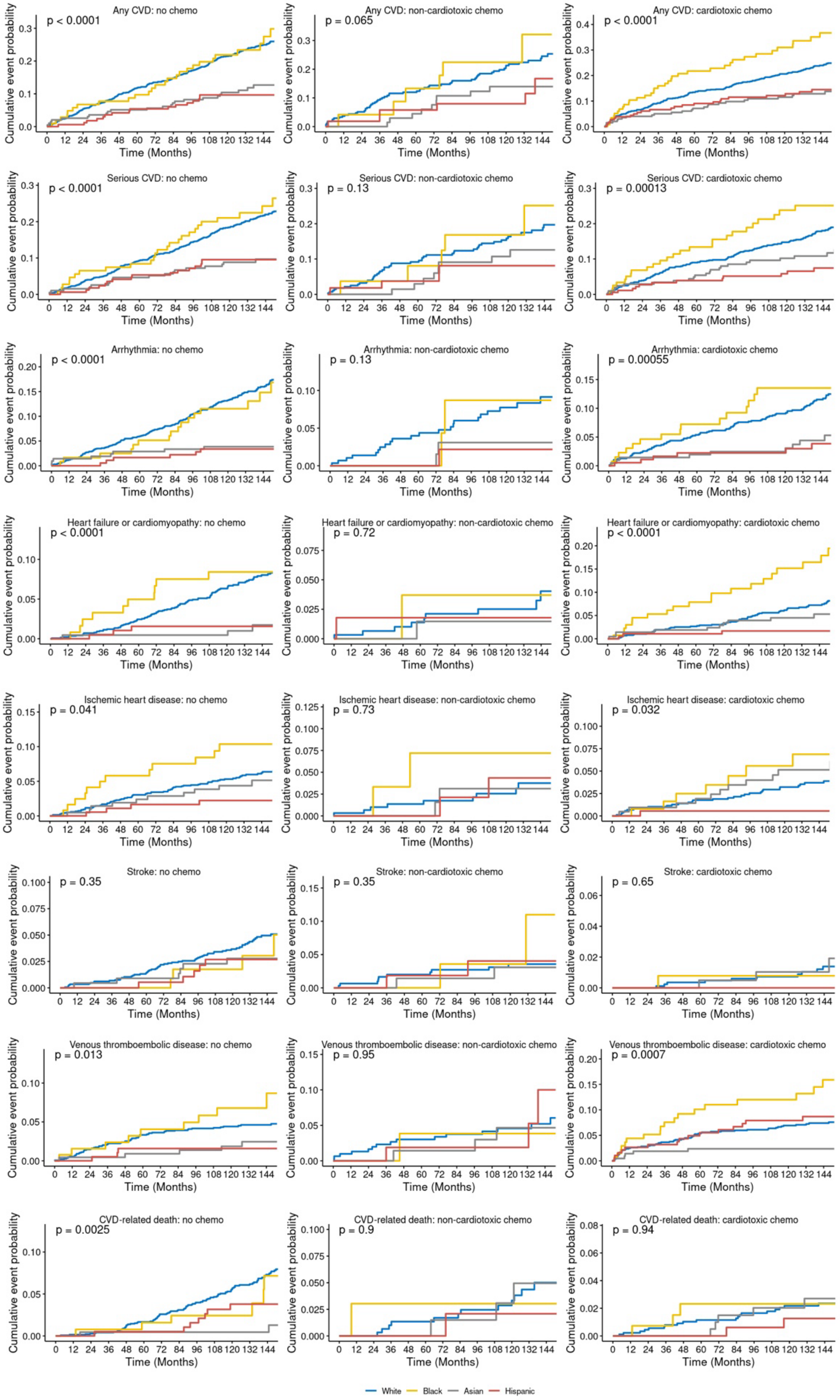
